# Effect of non-drug Intervention on Cognitive Function and Blood Sugar Control of Type 2 Diabetes patients with Mild Cognitive Impairment: A Meta-analysis Protocol

**DOI:** 10.1101/2024.09.19.24314026

**Authors:** Su Wenhao, Jia Hairong, Wang Yanru, Yang Luo, Li Xueling, Zhang Jiaqi, Wei Zhaoyang, Tsikwa Pepertual

**Affiliations:** School of Nursing, Zhejiang Chinese Medical University, Hangzhou, Zhejiang, China; School of Nursing, Hainan Vocational University of Science and Technology, Haikou, Hainan, China

**Keywords:** cognitive dysfunction, mild cognitive impairment, type 2 diabetes, non-drug intervention

## Abstract

This study aims to assess non-drug therapies’ safety and effectiveness on cognitive function and blood glucose control of type 2 diabetes with mild cognitive impairment (T2DM-MCI) patients in randomized controlled trials by meta-analysis, providing constructive evidence for non-drug treatment decision-making. The system review plan will strictly follow the Systematic Review and Meta-Analysis Protocols entry for reporting. PubMed, EMBASE, Cochrane Library, CINAHL, Web of Science, CNKI, WANGFANG Database, and SinoMed will be systematically searched with Chinese and English language restrictions, and all randomized controlled trials comparing non-drug treatment with usual care or no intervention or placebo to study cognitive impairment in T2DM-MCI patients will be included. We will also manually search for cited literature. Our primary outcomes are cognitive function and blood sugar control. The risk of bias will be assessed for all studies using the Cochrane risk-of-bias tool (RoB 2) for Systematic Review of Intervention-version 5.1.0. Where possible, meta-analysis using random-effects models will be performed, otherwise, a qualitative summary will be provided. We will adhere to the Preferred Reporting Items for Systematic Reviews and Meta-Analyses (PRISMA-P) guidelines. This meta-analysis compares the efficacy of non-drug interventions for mild cognitive impairment in type 2 diabetes mellitus. It will provide reliable evidence for patients, clinicians, and researchers.

The methodological protocol was in registered in the PROSPERO (CRD 42024496248).

## Background

By 2021, there were 529 million people with diabetes mellitus, and by 2050, that number is predicted to rise to 1.31 billion[1]. Among chronic noncommunicable illnesses, diabetes ranked fourth and contributed to 2 million deaths in 2019[2]. Furthermore, the incidence rate of mild cognitive impairment (MCI) is rising in tandem with the acceleration of the global aging process. Worldwide, the percentage of older persons with MCI who reside in nursing homes is around 21.2%[3], with the majority of older adults in China having MCI at a rate of approximately 15.5%, or 38.77 million[4].

Speech problems, memory loss, declines in executive function, and cognitive dysfunction are all considered symptoms of cognitive impairment in type 2 diabetes mellitus (T2DM)[5]. Diabetes also speeds up the pathological changes in the brain’s cognitive regions, leading to the deterioration of cognitive function and increasing the risk of dementia[6]. Studies reveal similarities in the pathophysiological mechanisms underlying diabetes and MCI[7].

Patients with diabetes have a risk of cognitive impairment that is more than double that of the general population[7], and those with type 2 diabetes have a likelihood of MCI that ranges from 30.7% to 70%[8-12]. Patients with type 2 diabetes have an incidence rate of dementia of 83/10000 person-years among those 60-64 years of age, and an incidence rate of more than 1000/10000 person-years among those 85 years of age and beyond[13]. The long-term nature of diabetes[14], inadequate blood glucose control[15], insulin resistance[16], etc., may all contribute to the cognitive impairment seen by T2DM patients.

Furthermore, cognitive dysfunction makes it harder to regulate blood sugar, which exacerbates diabetes[17]. A causal association has been shown between poor blood sugar control and the development of cognitive impairment. This relationship has a significant negative influence on the physical and mental health of patients, as well as increasing the financial and health burden of individuals with type 2 diabetes who also have mild cognitive impairment (T2DM-MCI). The vicious cycle between cognitive impairment and type 2 diabetes eventually developed.

Numerous investigations have verified that individuals with type 2 diabetes have a higher chance of cognitive dysfunction[5]. There are presently no effective medications accessible to treat MCI. MCI is a transitional stage from average cognitive decline to dementia[18]. Non-drug intervention is therefore at the top of the list. Researchers are progressively becoming interested in ways to put off or boost the development of MCI via the use of non-pharmacological measures[18].

While there have been some encouraging findings from non-drug intervention research on T2DM-MCI, there aren’t enough reliable meta-analyses combining high-caliber randomized controlled trials (RCT). To give evidence-based support for non-drug clinical treatment, our work attempts to synthesize the RCT studies of non-drug intervention for T2DM-MCI by systematic assessment.

### Objectives

This protocol for a systematic review and meta-analysis aims to compare various non-drug therapies to T2DM-MCI patients, and their efficacy and safety. We also aim to describe the advantages and shortcomings of different non-drug therapies for TDM-MCI patients diagnosed by MMSE, MoCA, or laboratory testing.

A comprehensive understanding of the current level of evidence in the literature will be useful for providing new ideas about the non-drug interventions of diabetic patients targeted and informing future research.

## Methods and analysis

This protocol is registered with the open access registry for systematic review protocols PROSPERO(CRD42024496248) and developed following the Preferred Reporting Items for Systematic Review and Meta-analyses for Protocols (PRISMA-P) 2015 checklist(S1 File)[19]. The planned systematic review will be reported according to the PRISMA 2020 statement[20]. S1 Fig. shows the picture chart of the protocol for the meta-analysis review steps.

### Eligibility criteria

A summary of eligibility criteria for Population, Intervention, Comparison, Outcome, and Study design (PICOS) (S2 File).

### Population Inclusion criteria

Trials that aimed to investigate people aged 60 years, with type 2 diabetes and mild cognitive impairment, will be included. Type 2 diabetes is defined as a group of metabolic diseases characterized by chronic hyperglycemia and insulin resistance, which can cause long-term damage to the brain, kidneys, nerves, etc[21].

### Exclusion criteria

Trials that investigated participants with suspected or confirmed psychiatric disease or neurological disorders (i.e., dementia, Alzheimer’s disease, type 1 diabetes, other unrelated endocrine diseases) were excluded. Trials that investigated efficacy of drug treatments and other surgical procedures (e.g., hypoglycemic drugs, pancreas surgery) will be excluded as well. We will also exclude studies with any of the following characteristics: The study reported that the control group’s plan was unclear; the intervention group’s treatment measure, frequency, and duration, outcomes were unclear; the data on cognitive function scores could not be extracted separately; documents not written in Chinese or English will be excluded.

### Interventions

According to the types of non-drug interventions, we performed previous search to classify the intervention measures and differentiate them into the list based on the mechanism. If an intervention cannot be distributed to a specific group, it will be included in a new group.

- Traditional Chinese exercise (Taichi, Qigong, etc.)
- Acupuncture
- Aerobic exercise(yoga, jogging)
- Resistance exercise
- Mindfulness(meditation, memory)
- Puzzle game
- Music therapy
- Mixed therapy
- Skillfulness game

### Comparator

The comparative group includes no intervention, placebo or sham intervention, and routine care. The routine nursing intervention group should indicate that participants have not received any other relevant interventions. If the study does not specify specific intervention measures, it will be included in the intervention-free group. The placebo group or sham intervention group should specify the type of placebo, otherwise, it will be excluded or included in the intervention group.

### Outcomes

Cognitive function and blood sugar levels will be considered if measured using a valid instrument.

### Cognitive function

We will preferentially extract data measured with Montreal Cognitive Assessment (MoCA) and Mini-Mental State Examination (MMSE). If the above rating table displays the intervention effect based on scores from different cognitive domains, we will extract data and compare them based on their cognitive domain division. We will prioritize extracting cognitive rating data for interventions lasting more than 6 months.

### Blood sugar control

We will prioritize extracting data on glycated hemoglobin, 2-hour postprandial blood glucose, and fasting blood glucose levels 6 months after intervention. In this review, the evaluation of multidimensional effects or interferences on blood glucose will not be considered.

### Study designs

RCT studies investigating at least one non-pharmacological intervention measure will be included in the study. Randomization can be performed at the individual or group level, and for crossover designs, only the data from the first trial period will be extracted to avoid other influencing factors.

### Search strategy

Search strategies will be conducted on the following: MEDLINE, Cochrane Library, Embase, Web of Science, CNKI, WANGFANG Database, and SinoMed.

The following keywords will be searched (randomized controlled trial*, RCT) AND (type 2 diabetes, diabete*, Non-Insulin-Dependent Diabetes Mellitus) AND (mild cognitive impairment).

Filters: last 5 years of the publication date, only RCT in Chinese and English will be considered.

### Study selection

After searching, the references will be exported to an EndNote file, and duplicates will be removed. Then, two independent reviewers (S. W. and L. X.) will screen titles and abstracts and will assess potential full texts. Those trials fulfilling our eligibility criteria will be included in the review. If necessary, authors will be contacted by email to clarify information. Three emails will be sent 5 days apart. If the authors do not answer, the study will be excluded, and the reasons will be reported in a flowchart. Between-reviewer discrepancies will be resolved by a third reviewer (W. Y.). Here is the literature collection process in (S2 Fig).

Fig. 2 PRISMA flow diagram of systematic review.

## Data extraction

Two independent reviewers (S. W. and L. X.) will extract characteristics and outcome data from included trials, and discrepancies will be resolved by a third reviewer (W.Y.).

Extracted data will include study design (i.e., parallel-group, crossover, RCT), source of participants, age, intervention details (e.g., types of treatment, intervention details, duration, frequency), and outcome data (including assessment used, timing, missing data details). For our outcomes of interest, we will extract from all groups: sample sizes, means, and standard deviations (SDs) or standard error, range score, interquartile range, and confidence interval. Short-term effects will be considered follow-ups up to 3 months after the baseline, and long-term effects will be considered follow-ups to 6 months after the baseline. If more than one time point is available within the same follow-up period, the one closer to the end of the intervention will be considered. Mean changes from baseline and theirs will be extracted if post-intervention scores are not available.

When trials include two or more arms comprehending frequency, or intensity of the same intervention, we will combine outcome data following the Cochrane recommendations[22]. In trials where SDs are not available, they will be imputed from the standard error, confidence interval, p-value, range values, interquartile interval, or other similar trials included, following the recommendations[22]. If SDs are not available, they will be imputed from the standard error, confidence interval, p-value, range values, and interquartile interval. When imputations are not possible, the authors will be contacted by email. If the authors do not respond, the study will be included in the review, but it will be excluded from the quantitative analysis. If data are missing due to participant dropout, we will use reported results for participants who completed the study.

Data extraction will be conducted by two independent reviewers (S.W. and L. X.) using previously prepared electronic forms. Discrepancies will be resolved by a third author (W. Y.). For crossover RCTs, we will only consider results from the first randomization period to avoid carryover effects. The data will be extracted following the recommendations in the Cochrane Handbook[22].

### Risk-of-bias assessment using the Cochrane risk-of-bias tool (RoB 2)

Two independent trained reviewers (S. W. and L. X.) will assess the methodological quality of the included trials. The risk of bias will be assessed for all trials using the revised Cochrane risk-of-bias (RoB) tool 2.0[23]. The following five domains will be assessed: (1) bias arising from the randomization process, (2) bias due to deviations from intended interventions, (3) bias due to missing outcome data, (4) bias in the measurement of the outcome, and (5) bias in the selection of the reported result.

We will use the algorithms described in the instrument for classification of each domain as follows: (1) low risk of bias, (2) some concerns, and (3) high risk of bias. The judgment of the overall risk of bias of the included trial will follow the rule: (1) low risk of bias, low risk of bias for all domains; (2) some concerns, some concerns for at least one domain but no high risk of bias in any domain; and (3) high risk of bias, high risk of bias in at least one domain or have some concerns for multiple domains in a way that substantially lowers confidence in the result.

### Strategy for data synthesis and analysis

Basic information about literature, baseline information, and experimental results will be extracted, including article title, author, year of publication, study location, study size, follow-up duration, age at baseline, sex distribution, duration of T2DM-MCI, and study outcomes of Montreal Cognitive Assessment scores or Mini-mental State Examination score, fasting glucose, HbA1c, ADL, 2hPG. The odds ratio and 95% confidence interval (CI) will be calculated to assess the association between non-drug intervention and T2DM-MCI. If more than 3 RCTs are included, the state software will be used for meta-analysis. According to the Cochrane Handbook[20], I^2^ is less than 20% with fixed effect model, 20% to 50% with random effect model, and more than 50% with subgroup analysis. When there are more than 9 studies, a sensitivity analysis will be conducted.

A sensitivity analysis will also be carried out if only a small number of RCT studies are included. In this analysis, one study at a time is eliminated and the remaining studies are examined to determine whether the removal of one study will significantly alter the results. When sensitivity analysis is ineffective, a thorough assessment of the pertinent literature will be conducted. To better understand the current status of the field and recent advancements in the study of T2DM-MCI, we will undertake a literature review on T2DM-MCI studies. And draw a conclusion on the present development, highlighting both its advantages and disadvantages.

### Assessing the quality of evidence

Two reviewers will independently evaluate the quality of the evidence for blood sugar (fasting glucose, 2hPG) and cognitive performance (MCA, MMSE). The Grading of Recommendations Assessment, Development, and Evaluation (GRADE)[24] will be employed to complete the task. Five dimensions need to be considered: publication bias, indirectness, inconsistency, risk of bias, and imprecision. This will provide a high, moderate, low, or very low certainty of the evidence assessment, which will be displayed in a summary of findings table.

## Discussion

The study wants to provide the protocol that will be used in the systematic review and meta-analysis to evaluate the role of non-drug interventions in mild cognitive impairment of T2DM patients and analyze the mechanism as much as possible. The significance of a review and meta-analysis is to evaluate its effect in a larger sample and to summarize the current research results for further advice on clinical research, which will have a positive significance in protecting the cognitive dysfunction of diabetic patients. Since this research methodology produces a basis for public health and clinical decision making it is crucial to publish specific Protocols like the present paper not only to minimize bias risks for hereby author group but mostly to help future research serving as an example to further meta-analysis with similar comparators and outcomes. We will use Revman 5.4 software to analyze the bias sharing included in the study, including each result and adverse reaction in each trial.

## Data Availability

No datasets were generated or analysed during the current study. All relevant data from this study will be made available upon study completion.

## Author Contributions

Conceptualization: Su Wenhao, Wang Yanru

Data curation: Su Wenhao, Jia Hairong

Formal analysis: Jia Hairong, Su Wenhao

Investigation: Su Wenhao, Zhang Jiaqi, Wei Zhaoyang

Methodology: Su Wenhao, Li Xueling, Wang Yanru

Supervision: Wang Yanru

Writing-original draft: Su Wenhao, Jia Hairong, Yang Luo, Tsikwa Pepertual

Writing-review & editing: Su Wenhao, Wang Yanru

## Supporting Information

S1 Fig. Chart of the protocol for the meta-analysis review steps.

(TIF)

S2 Fig. PRISMA flow diagram of systematic review.

(TIF)

S1 File. PRISMA-P 2015 checklist.

(PDF)

S2 File. A summary of eligibility criteria for Population, Intervention, Comparison, Outcome, and Study design (PICOS)

(PDF)

### Acknowledgements

No applicable

## Author Contributions

Conceptualization: Su Wenhao, Wang Yanru

Formal analysis: Jia Hairong, Su Wenhao

Investigation: Su Wenhao, Zhang Jiaqi, Wei Zhaoyang,

Methodology: Su Wenhao, Li Xueling, Wang Yanru

Supervision: Wang Yanru

Writing-original draft: Su Wenhao, Jia Hairong, Yang Luo, Tsikwa Pepertual

Writing-review & editing: Su Wenhao, Wang Yanru

## Funding

The authors declared no specific for this research from any funding agency in the public, commercial, or not-for-profit sectors.

## Declarations

Ethic approval and consent to participate Not applicable

## Consent for publication

Not applicable

## Competing for publication

Not applicable

## Competing interests

The authors declare that they have no competing interests.

## Notes

### Competing Interest Statement

The authors have declared no competing interest.

### Funding Statement

The author(s) received no specific funding for this work.

